# Nationwide survey of municipal policies and COVID-19 vaccination uptake among older adults in Japan during 2024–2025

**DOI:** 10.1101/2025.07.17.25330544

**Authors:** Masaki Machida, Akira Nomachi, Kosuke Tanabe, Shuhei Ito, Tomoki Nakaya, Takashi Matono

## Abstract

Many countries continue to recommend COVID-19 vaccination, primarily to older adults. Following the transition of the national immunization program for COVID-19 vaccination in Japan in April 2024, implementation was delegated to municipalities. However, vaccination uptake and its associated factors, particularly municipal-level policies, remain unclear. This ecological study clarifies the factor associated with COVID-19 vaccination uptake among older adults in Japan during 2024–2025 by analyzing 1,740 municipalities. Municipal vaccination uptake among individuals aged ≥65 years was estimated from vaccine distribution data for the period. We examined the associations of uptake with municipal policies and sociodemographic factors using negative binomial regression, and incidence rate ratios (IRRs) and 95% confidence intervals (CIs) were calculated. The mean estimated vaccination uptake across Japan was 17.9%. Individual notification via mailed vaccine screening questionnaires had the largest effect, with higher uptake (IRR 1.57; 95% CI, 1.37–1.80). Low out-of-pocket expenses were also associated with higher uptake (IRR 1.31; 95% CI, 1.12–1.52 for ≤2,000 JPY, Reference: ≥3,001 JPY). A higher proportion of those aged ≥75 (IRR 1.03; 95% CI, 1.01–1.05) and a higher number of physicians (IRR 1.03; 95% CI, 1.00–1.06) were positively associated with uptake as well. Conversely, the area deprivation index was associated with lower uptake (IRR 0.87; 95% CI, 0.76–0.98). Proactive municipal strategies (individual notifications and subsidies to reduce out-of-pocket expenses) were significantly associated with increased vaccination uptake. To ensure equitable access and mitigate disparities, future vaccination campaigns should incorporate outreach measures to address socioeconomic barriers.

## Introduction

During the COVID-19 pandemic, various policies have been implemented across countries to increase vaccination uptake, including free vaccination and the establishment of mass vaccination sites. Since the transition to the Omicron phase, the pathogenicity of SARS-CoV-2 has not changed substantially,^1,2^ and many countries continue to recommend COVID-19 vaccination, mainly for older adults.^3^

In Japan, the Ministry of Health, Labour and Welfare has conducted special temporary vaccinations under the Immunization Act in response to urgent public health needs to prevent the spread of COVID-19. This has led to the implementation of unprecedented policies, such as providing COVID-19 vaccinations free of charge, establishing special vaccination sites, and developing a dedicated vaccine distribution system. As of March 2022, approximately 80% of individuals had received two doses of the vaccine.^4^ The special temporary vaccination program, under which eligible individuals were obligated to make efforts to receive the vaccine, ended on March 31, 2024. Therefore, since April 2024, COVID-19 vaccinations have been administered as routine vaccinations to prevent severe illness, and the disease has been reclassified as a category B disease, similar to influenza, under the Immunization Act.^5^ Regarding routine vaccines for category B diseases, municipalities, rather than the national government, are tasked with implementing public awareness activities, including mailing vouchers for vaccines, managing records, and administering potential subsidies. Vaccine subsidies are limited to individuals aged 65 years and older and those aged 60–64 years with certain underlying medical conditions.^6^

Because policies such as COVID-19 vaccine subsidies and vaccination promotion activities vary across municipalities, regional differences in vaccination uptake may reflect variations in municipal policies. However, as few studies have examined the association between vaccination uptake and municipal policies in Japan,^7^ this study aimed to clarify the association between COVID-19 vaccination uptake and its associated factors, including sociodemographic factors and municipal policies, among older adults in Japan during 2024–2025.

## Materials and Methods

### Data sources

This exploratory study adopted an ecological design. As COVID-19 vaccinations are mainly conducted for those aged 65 years and older, this study included the vaccination uptake among those individuals. All subsidy periods began on October 1, 2024, while the end date for these periods varied by municipality. The final possible end date was March 31, 2025, for a maximum of 182 days. In addition, as official data on COVID-19 vaccine uptake had not been made publicly available following the start of routine vaccination in Japan at the time of the survey, the COVID-19 vaccination uptake was estimated using Pfizer’s COVID-19 vaccine production, inventory, and return numbers, along with municipal population data. During this period, Pfizer’s COVID-19 vaccine was widely administered in Japan,^8^ accounting for a substantial proportion of the total vaccinations.

Sociodemographic factors previously reported to be associated with other vaccination uptake, along with municipal policy, were evaluated as independent variables.^9^ Data on population and sociodemographic factors by municipality were obtained from e-Stat, a portal site for Japanese Government Statistics.^10^ Information on municipal policies were obtained from official municipal websites that made this information available. If information on COVID-19 vaccination for a particular municipality were not publicly accessible online, the municipal vaccination officer was contacted by telephone. We defined a municipal policy as “not implemented” when no information of its implementation could be confirmed on the website or by telephone. In cases where the number of vaccination sites and subsidy duration could not be confirmed on the website or by telephone, the data were marked as missing. Municipalities with no persons eligible for the COVID-19 vaccine were excluded from the analysis because of missing population data. Ethical review was not required because the study used only publicly available data. Sales data for the vaccinations that were collected for this study represented regional-level summaries, and did not include any personally identifiable information.

### Dependent variables

The estimated number of COVID-19 vaccine recipients was calculated using data on COVID-19 vaccine production, inventory, and returns by municipality during the 2024–2025 season in Japan. The population aged 65 years and older as of January 1, 2024, was extracted from the 2024 Basic Resident Registration.^10^ COVID-19 vaccination uptake was defined as the estimated number of COVID-19 vaccine recipients divided by the population aged 65 years and older, as of January 1, 2024. If the uptake calculated using this method exceeded 100%, any uptake exceeding 100% was converted to 100%.

### Independent variable: sociodemographic factors

Population density was defined as the number of people per square kilometer (number/km^2^), and the number of employed individuals aged 65 years and older was extracted from the 2020 census of Japan, which is the most recent data source available. Population density was transformed using its natural logarithm because of the right-skewed distribution. The regional classification was based on the most commonly used geographic divisions in Japan: Hokkaido, Tohoku, Kanto, Chubu, Kinki, Chugoku, Shikoku, and Kyushu. The proportion of workers aged 65 years and older was calculated by dividing the number of employed individuals aged 65 years and older by the total population of the same age.

The area deprivation index (ADI) derived from the 2020 census was used as an indicator of municipality-level socioeconomic status. This composite indicator is the weighted sum of census variables related to poverty. A higher ADI score indicates greater deprivation in the municipality. The details of the ADI calculation are provided elsewhere.^11^ The data on the population aged 65 and older and aged 75 and older as of January 1, 2024 were extracted from the 2024 Basic Resident Registration. The percentage of individuals aged 75 and older was calculated by dividing the population aged 75 and older by the population aged 65 and older. The number of physicians was obtained from the Statistics of Physicians, Dentists and Pharmacists for 2022. ^12^ The number of physicians per 1,000 population was calculated using population data from the 2020 census.

### Independent variable: municipal policies

The following municipal policies were investigated to assess the implementation of vaccination-related information dissemination (implemented/not implemented): mailing vaccine screening questionnaires, notifications via communication apps (e.g., LINE®), notification on social media (e.g., X®), postings on official municipal websites, creating campaign posters, publication in public relations magazines, reminders about subsidy deadlines, and making campaign flyers. In Japan, everyone who receives a vaccination must fill out a vaccine screening questionnaire to confirm their health status, vaccination records, and signatures of consent. The number of vaccination sites per square kilometer was calculated by dividing the number of vaccination sites by the inhabitable land area, as reported in the statistical observations of municipalities 2024.^10^ The number of vaccination sites per square kilometer was classified into two groups based on the median value: less than 0.3 sites per km^2^ and 0.3 or more. Regarding financial subsidies, the methods for determining subsidy amounts varied among municipalities and generally followed one of these three patterns: (1) a fixed out-of-pocket expense for vaccine recipients; (2) a specified subsidy amount provided by the municipalities; or (3) a specified percentage of the total vaccination cost to be paid by the recipient. In this study, out-of-pocket expenses were used as the independent variable. Therefore, in cases (2) and (3), where the exact out-of-pocket amount was not stated, the cost was based on a standard vaccination cost of JPY 15,300 (approximately $105 USD), which was calculated as the amount to be paid by an individual from the subsidies allocated by each municipality. We categorized out-of-pocket expenses based on their distribution, which was highly concentrated in the JPY 2,000–3,000 range. For descriptive purposes, we established four categories: JPY 0, 1–2,000, 2,001–3,000, and ≥3,001.

However, for the multivariable regression model, we combined the JPY 0 and 1–2,000 categories. This decision was made to ensure sufficient statistical power and model stability given the relatively small number of municipalities (n=49) with no out-of-pocket expenses. Therefore, the final independent variable for the multivariate analysis consisted of three levels: JPY 0–2,000, 2,001–3,000, and ≥3,001, with the highest-cost category serving as a reference. We also evaluated the policies related to the municipal subsidy period. The maximum possible subsidy duration was 182 days (October 1, 2024, to March 31, 2025). Although some municipalities set shorter periods initially, the Ministry of Health, Labour and Welfare encouraged an extension of these periods. To examine whether encouragement from the government had an impact on the vaccination uptake, the extension of the subsidy period was also included as a variable. Reflecting on this context, we created a composite categorical variable, “subsidy duration and extension,” that combined two dimensions: (1) whether the municipality extended its subsidy period and (2) whether the final duration covered the full 182 days. This resulted in a four-category variable: (1) full period (182 days) subsidy without extension, (2) shorter than the full period without extension, (3) full period (182 days) following extension, and (4) shorter than the full period following extension.

### Statistical analysis

First, we calculated descriptive statistics for all variables. For continuous variables, the mean, standard deviation (SD), range, and interquartile range (IQR) were calculated. For categorical variables, the number (n) and proportion (%) of municipalities in each category were calculated. We also calculated the mean COVID-19 vaccination uptake for each category of municipal policy variables. To visualize the geographic distribution of COVID-19 vaccination uptake across Japan, a choropleth map was created at the municipal level. Furthermore, to illustrate the association between the key policy variable of out-of-pocket expenses and vaccination uptake, a box-and-whisker plot was generated to compare the distribution of uptake across the four cost categories: JPY 0, 1–2,000 2,001–3,000, and ≥3,001. Finally, to identify the factors associated with COVID-19 vaccination uptake, a negative binomial regression analysis was performed. The dependent variable was the estimated number of COVID-19 recipients. The offset term was the population aged 65 years and older as of January 1, 2024. The independent variables were the following: mailing vaccine screening questionnaires, notifications via LINE^®^, notification on X^®^, postings on official municipal websites, creating campaign posters, publication in public relations magazines, reminders about subsidy deadlines, making campaign flyers, out-of-pocket expenses, subsidy duration and extension, number of vaccination sites, population density, proportion of aged 75 and older, proportion of workers aged 65 and older, ADI, number of physicians per 1,000 population, and region. To address missing data in the explanatory variables, we applied a maximum likelihood estimation under the assumption that the data were missing at random. This model-based approach allowed consistent estimates by leveraging all available data. Incidence rate ratios (IRR) were calculated for each indicator. The significance level used for statistical testing was 0.05, and the confidence level for the interval estimation was 95% on both sides. SAS 9.4 (SAS Institute Inc., Cary, NC, USA) was used to perform the statistical analyses.

## Results

### Characteristics of municipalities

The number of municipalities conducting COVID-19 vaccination was 1,741. Due to the exclusion of one municipality with no eligible people for the COVID-19 vaccine, the final dataset used for the analysis included 1,740 municipalities. The basic characteristics of these municipalities are listed in Table 1. The mean out-of-pocket expense for vaccination was JPY 2,727 (SD 927.7), and the mean proportion of aged 75 and older among those aged 65 and older was 55.4% (SD 3.8).

**Table 1.** Basic statistics of each indicator by municipality (n=1,740)

| Indicators (units) | Mean (SD) | Range (min, max) | IQR (Q1–Q3) |
| --- | --- | --- | --- |
| Population density <sup>a</sup> | 5.3 (1.9) | 0.3, 10.1 | 4.0–6.6 |
| Proportion of aged 75 and older (%) <sup>b</sup> | 55.4 (3.8) | 39.5, 70.8 | 53.1–57.8 |
| Proportion of workers aged 65 and older (%) | 27.3 (6.2) | 0.7, 80.4 | 23.3–30.3 |
| Area deprivation index <sup>c</sup> | 6.1 (0.7) | 3.6, 9.6 | 5.7–6.6 |
| Number of physicians per 1,000 population | 1.7 (1.9) | 0.1, 21.7 | 0.8–2.1 |
| Out-of-pocket expenses (JPY) <sup>d</sup> | 2727 (927.7) | 0, 7200 | 2100–3300 |
| Duration of subsidy (days) <sup>e</sup> | 160.9 (28) | 51, 182 | 123–182 |
| Number of vaccination sites per square kilometer <sup>f</sup> | 1.0 (2.0) | 0, 26.9 | 0.1–0.9 |

IQR: interquartile range; SD: standard deviation
<sup>a</sup> Population density, defined as the number of people per square kilometer of land area (number/km<sup>2</sup>), was subjected to a logarithmic transformation using the natural logarithm, owing to its right-skewed distribution.
<sup>b</sup> The number of people aged 75 and older divided by the number of people aged 65 and older, described in percentage terms.
<sup>c</sup> The area deprivation index derived from the 2020 census results was used as an indicator of socioeconomic status at the municipal level. This composite indicator comprises the weighted sum of poverty-related census variables.
<sup>d</sup> In cases where the amount to be paid by the patient was not specified, the estimated amount was calculated based on the standard cost of JPY 15,300 for vaccinations that were not covered by routine vaccination.
<sup>e</sup> Each municipality sets its own subsidy period, and the number of subsidy days is indicated. Although all subsidy periods began on October 1, 2024, the end date for these periods varied by municipality. The final possible end date was March 31, 2025. The maximum number of days eligible for subsidies is 182 days.
<sup>f</sup> Dividing the number of medical institutions administering COVID-19 vaccines by the inhabitable land area

### COVID-19 vaccination uptake

The estimated COVID-19 vaccination uptake across Japan for those aged 65 and older was 17.9%, and the mean COVID-19 vaccination uptake by municipality was 16.6%. Figure 1 illustrates the geographic distribution of the vaccination uptake, which reveals considerable variations across municipalities. The number of municipalities and the vaccination uptake for each category of out-of-pocket expenses are as follows, JPY 0: 49 municipalities and 22.2%, JPY 1–2,000: 377 municipalities and 18.4%, JPY 2,001–3,000: 757 municipalities and 16.6%, and ≥JPY 3,001: 557 municipalities and 14.7% (Figure 2).

**Figure 1.**
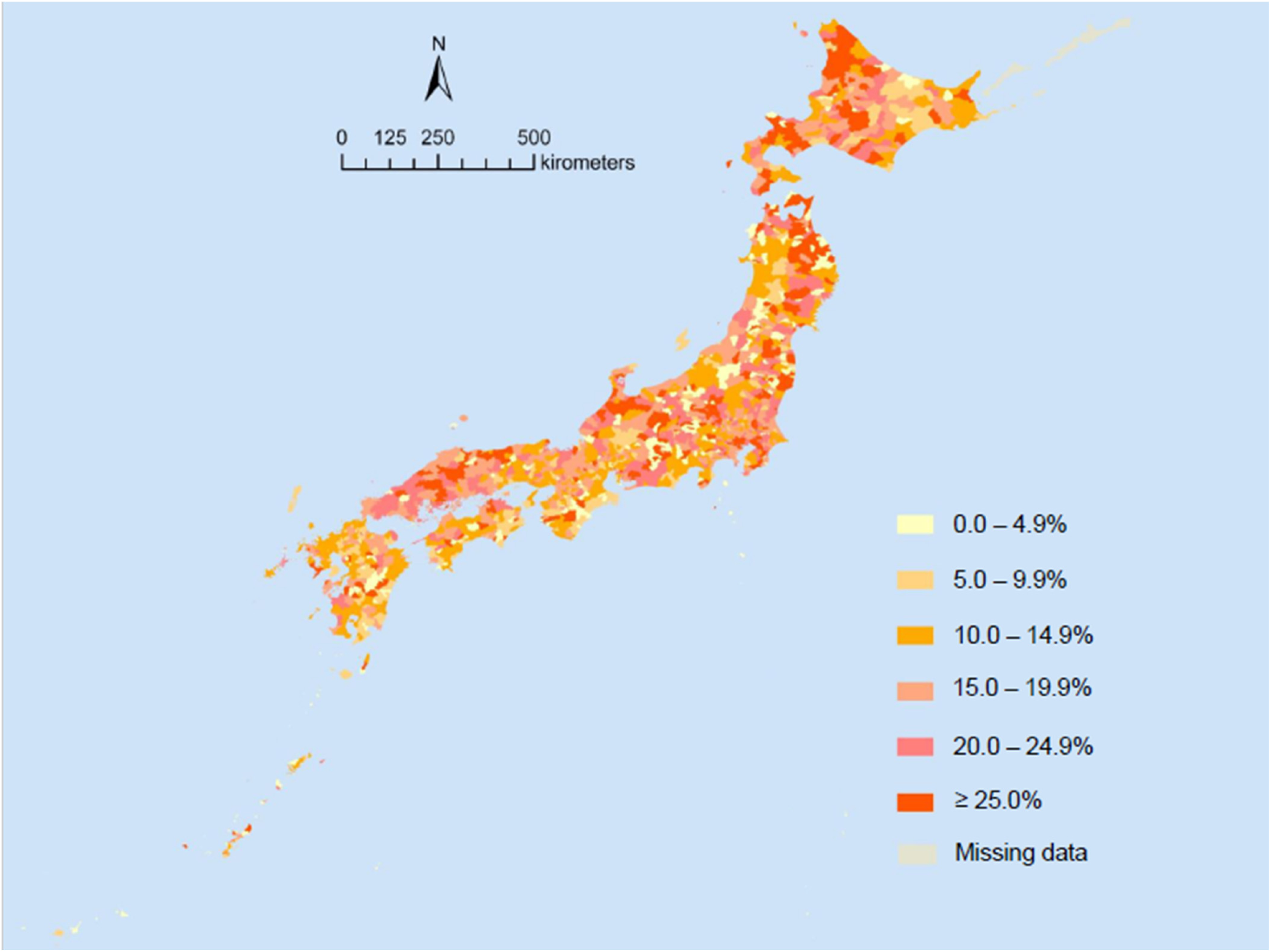
COVID-19 vaccination uptake by municipality in Japan

**Figure 2.**
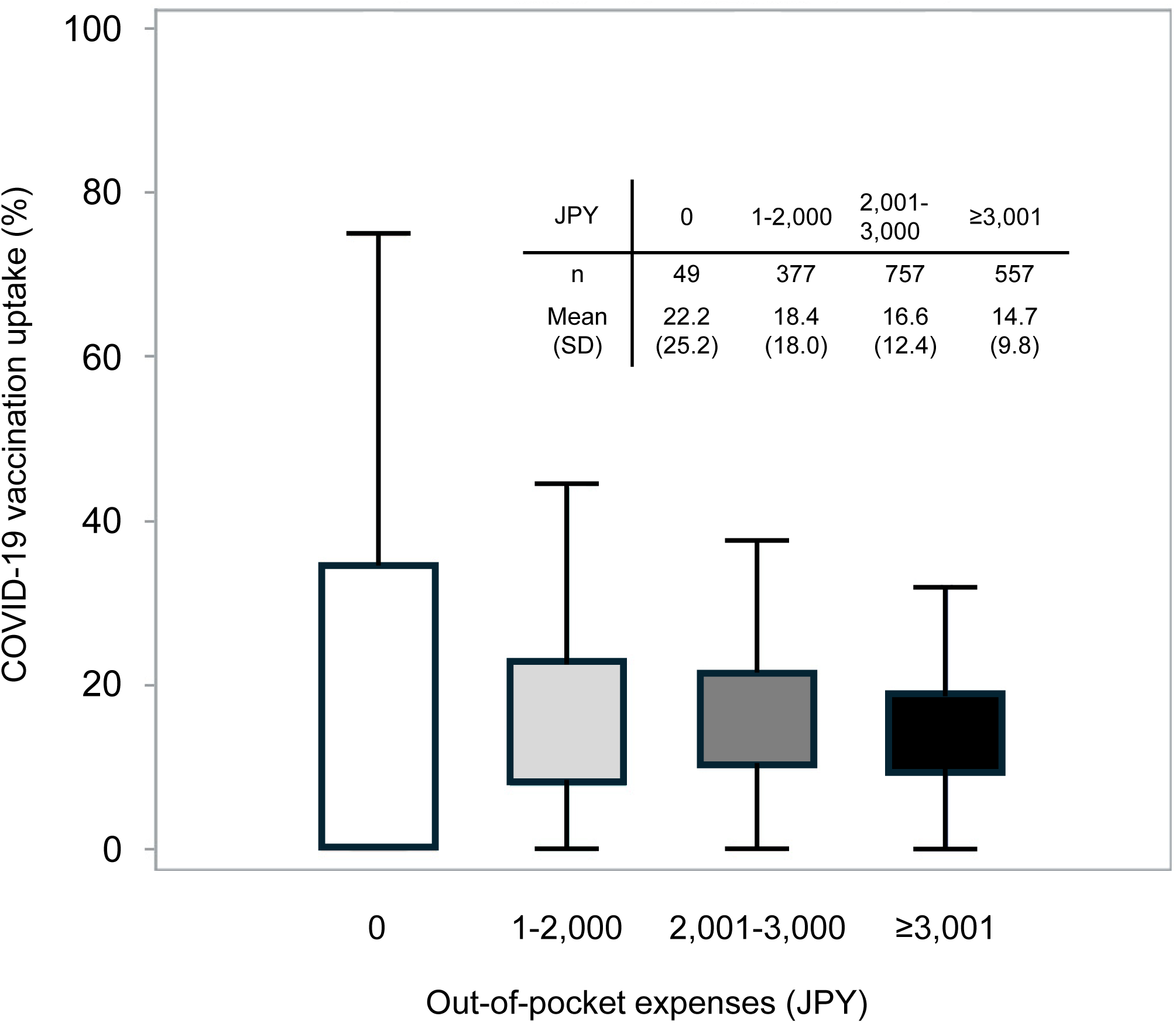
COVID-19 vaccination uptake by category of out-of-pocket expenses

Table 2 shows the implementation proportion of municipal policies and COVID-19 vaccination uptake by category. Many municipalities posted on official municipal websites and published in public relations magazines (78.7% and 83.7%, respectively). Meanwhile, vaccine screening questionnaires were mailed in 21.6% of municipalities.

**Table 2.**
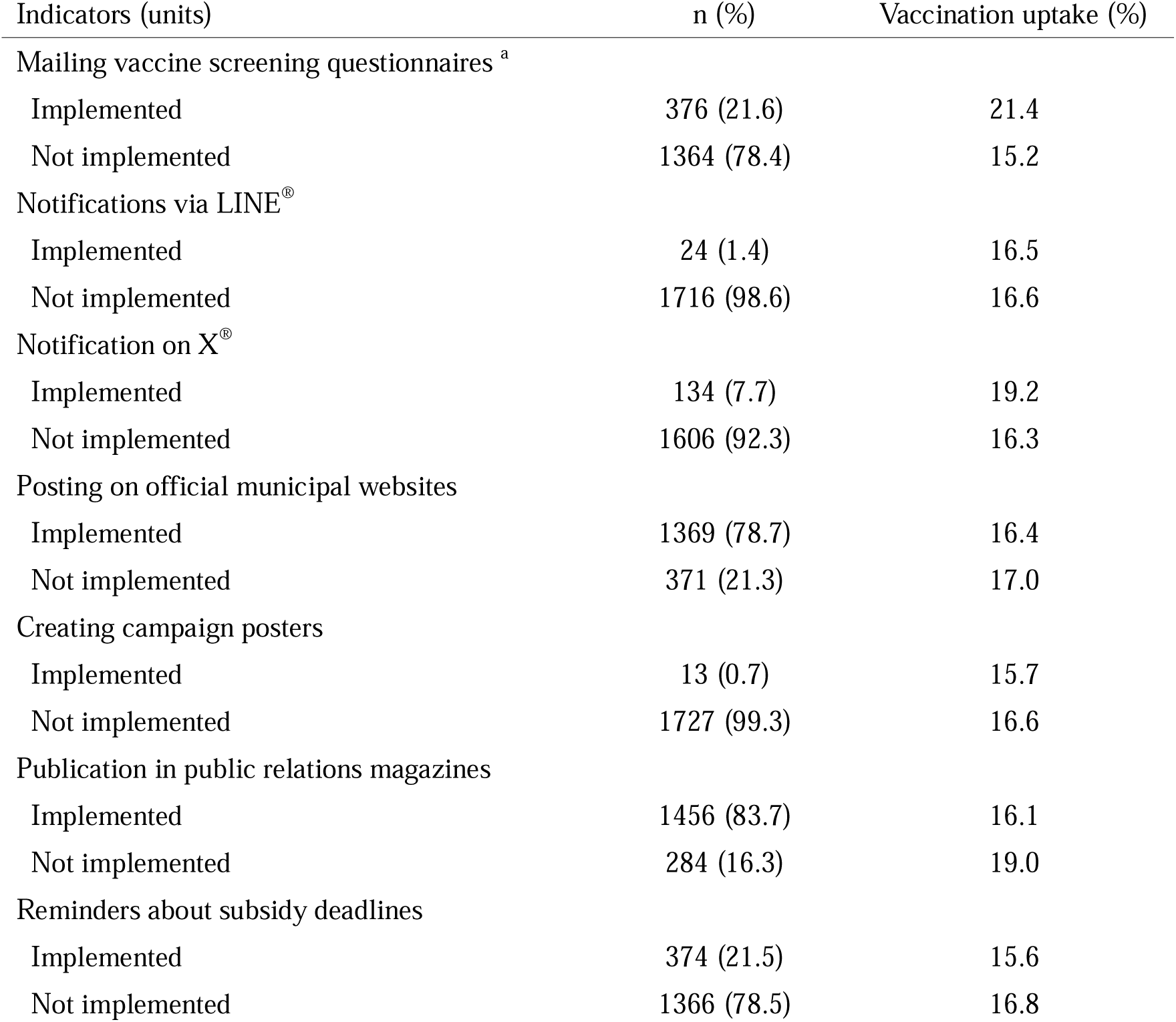
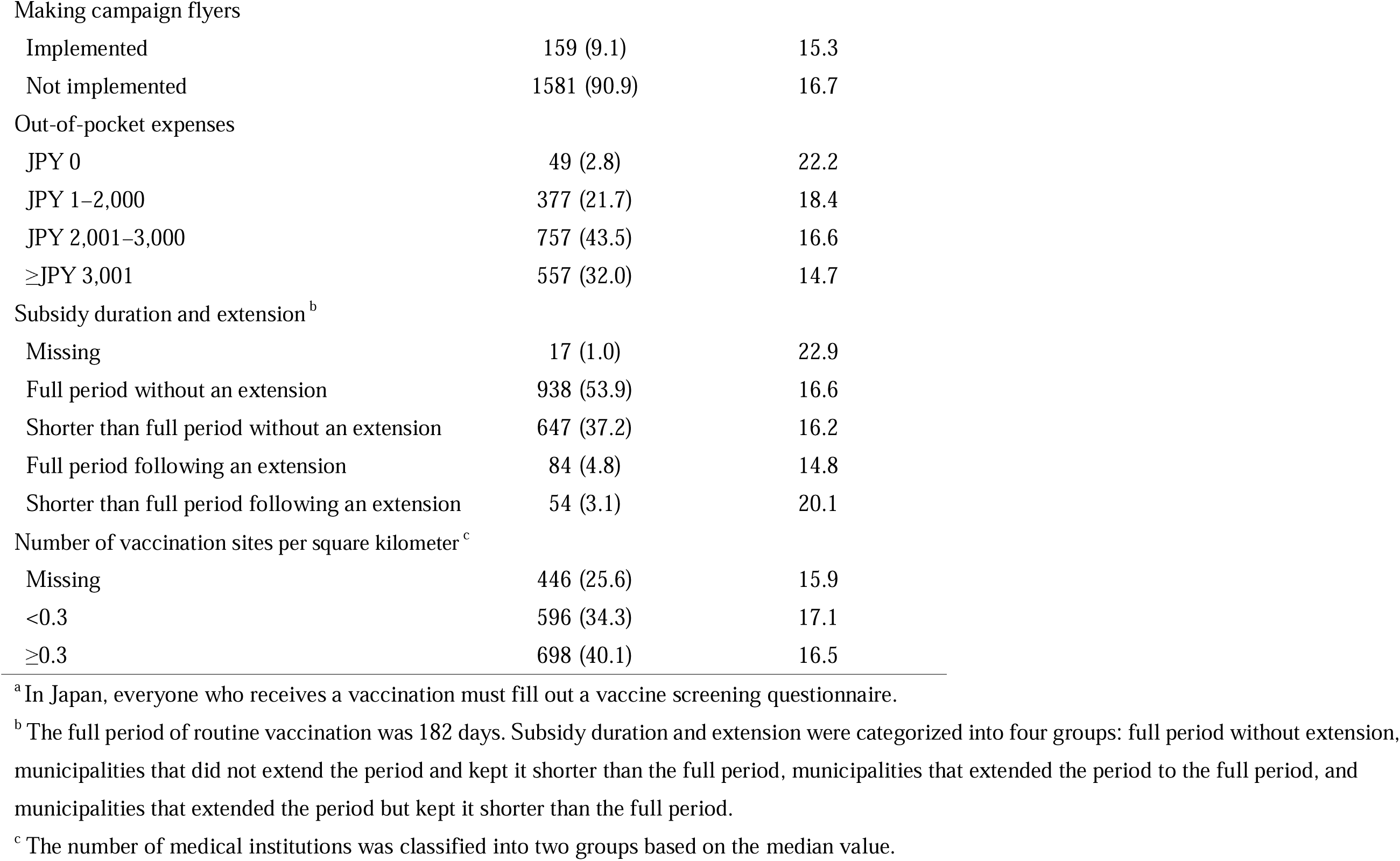
Proportion of municipal policies implemented and COVID-19 vaccination uptake by category.

### Factors associated with vaccination uptake in the negative binomial regression analysis

The results of the negative binomial regression analysis are shown in Figure 3. Among the evaluated municipal policies, mailing vaccine screening questionnaires showed the largest effect estimate for its association with higher uptake (IRR 1.57; 95% CI, 1.37–1.80). Lower out-of-pocket expenses also remained significantly associated with higher uptake compared to the reference group (≥JPY 3,001): the IRR was 1.31 (95% CI, 1.12–1.52) for expenses of JPY 0–2,000 and 1.15 (95% CI, 1.01–1.30) for expenses of JPY 2,001–3,000. Regarding sociodemographic factors, the proportion of those aged 75 and older (IRR 1.03; 95% CI, 1.01–1.05) and number of physicians per 1,000 population (IRR 1.03; 95% CI, 1.00–1.06) were positively associated with uptake. Conversely, ADI (IRR 0.87; 95% CI, 0.76–0.98) and the proportion of workers aged 65 and older (IRR 0.98; 95% CI, 0.97–0.99) were negatively associated with uptake. Most other informational campaigns, such as using notification via LINE^®^ or notification on X^®^, were not significantly associated with uptake in the adjusted model.

**Figure 3.**
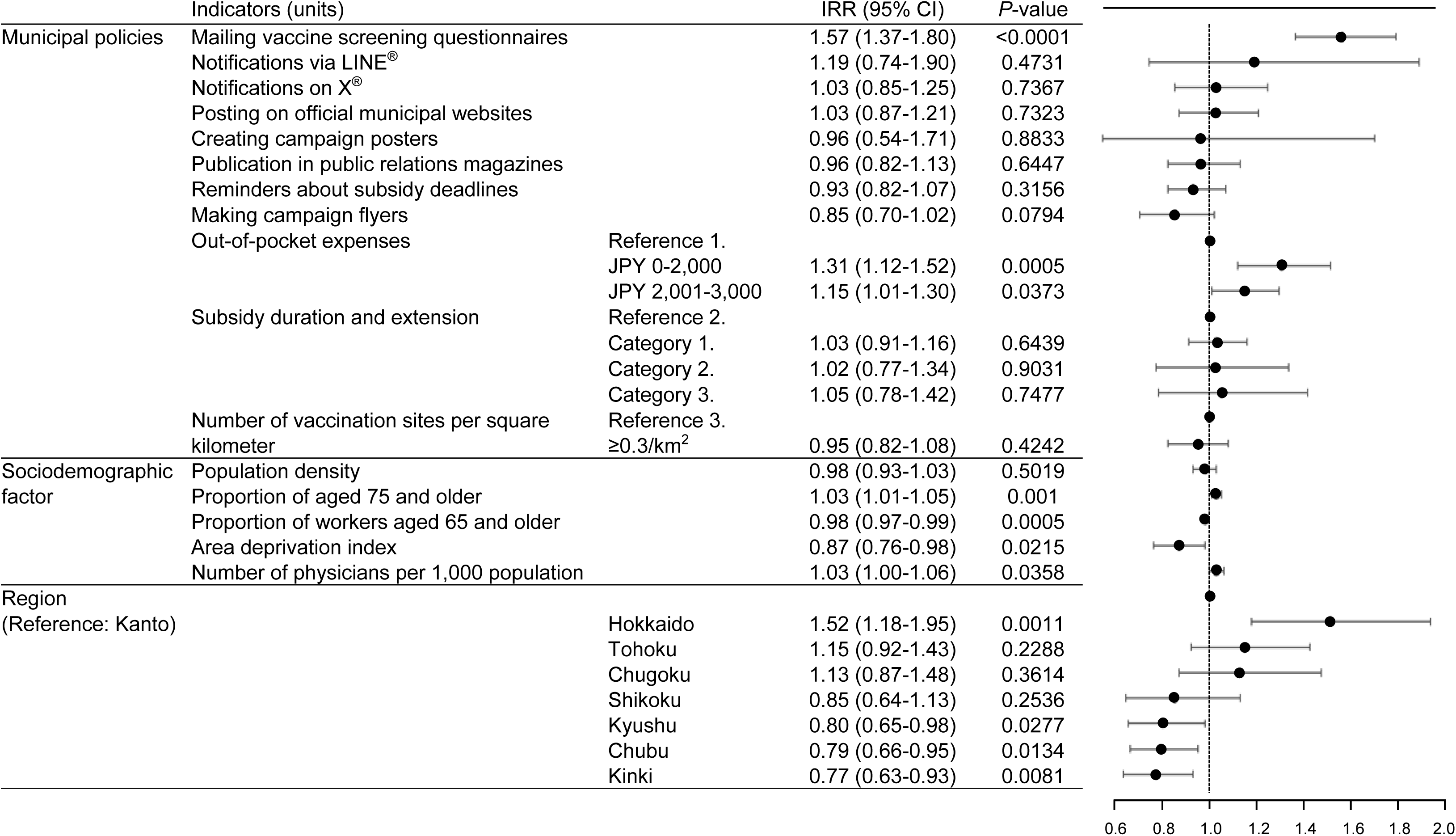
Factors associated with COVID-19 vaccination uptake Category 1. Shorter than full period without extension, Category 2. Full period following extension, Category 3. Shorter than full period following extension, Reference 1. ≥JPY 3,001, Reference 2. Full period without extension, Reference 3. <0.3/km^2^ IRR: Incidence Rate Ratio

## Discussion

In this ecological study conducted during the inaugural 2024–2025 season of Japan’s immunization program for COVID-19 routine vaccination, municipal-level policies, particularly direct communication and financial support, were significantly associated with COVID-19 vaccination uptake among older adults. To the best of our knowledge, this is the first study to clarify such associations for Japan’s new vaccination program. Specifically, mailing vaccine screening questionnaires was associated with the largest increase in uptake among the municipal policies, and a dose–response relationship was observed for lower out-of-pocket expenses. These findings underscore the important role of municipal strategies in maintaining vaccination uptake even after national mandates and recommendations have weakened, and they provide evidence for designing future public health interventions. The estimated national vaccination uptake was 17.9% across Japan, although vaccination uptake varied greatly depending on municipality. The COVID-19 vaccination uptake for special temporary vaccination was 54.4% among older adults in the 2023–2024 season, placing Japan in a challenging position when viewed in an international context.^13^ The lower uptake in the 2024–2025 season was likely attributable to a confluence of factors following the transition of the program. The special temporary vaccination programs for the COVID-19 vaccine include various policies, such as proactive recommendations by the government,^6,14^ free vaccinations for all older adults, and large-scale special vaccination sites. However, these were not included in the routine vaccination program in the 2024–2025 season. The vaccination uptake in the 2024–2025 season (17.9%) was lower than that reported in the United Kingdom^15^ (59.3%) and the United States^16^ (44.1%) for individuals aged 65 years and older. It is particularly noteworthy that Japan’s uptake declined significantly from the previous season’s (2023–2024) proportion of 54.4%,^13^ despite the vaccination uptake for individuals aged 65 years and older in the 2023–2024 season being higher than that in the United States (37.7%).^17^ While the US has maintained vaccination access through pharmacies and drugstores,^18^ Japan needs to implement effective strategies to increase the vaccination uptake.

This study demonstrated that lower out-of-pocket expenses are associated with a higher COVID-19 vaccination uptake. This association is consistent with previous studies on pneumococcal vaccines in Japan.^7,19^ In the 2024–2025 season, a special national fund limited the maximum out-of-pocket expenses to approximately JPY 7,000.^20^ With the addition of municipal subsidies, the mean cost of the COVID-19 vaccine was JPY 2,727. Our results suggest that an increase in out-of-pocket expenses can lead to a further decline in vaccination uptake, highlighting the importance of financial accessibility.

Beyond financial support, our findings underscored the impact of individual notifications. Specifically, mailing vaccine screening questionnaires had the strongest association with COVID-19 uptake and the highest IRR (1.57). An IRR of 1.57 indicates that the vaccination uptake was estimated to be 1.57 times higher in municipalities that mailed the questionnaire than in those that did not. Meanwhile, this study found no statistically significant association between vaccination uptake and mass marketing policies such as providing information on a public platform that is not individualized (e.g., notifications via X®, posting on official municipal websites, and publication in public relations magazines). This finding resonates with previous studies on pneumococcal vaccines in Japan, where a direct approach was associated with an increased uptake, while broad awareness campaigns such as providing information on websites, and distributing pamphlets had little effect on the vaccination uptake.^21^ This suggests that direct individual approaches such as mailing vaccine screening questionnaires may be a more impactful strategy for municipalities than broad awareness campaigns, particularly among older adults. However, given the ecological study design and the difficulty of perfectly measuring the implementation and reach of public relations activities, our study may have been unable to detect a modest but meaningful effect of mass marketing. These campaigns might play a foundational role in building public trust and supporting direct approaches, a possibility that requires the elucidation of different study designs. A practical application of this individual notification strategy is its integration with other routine vaccinations, such as the annual influenza vaccination program. Simultaneous administration of COVID-19 and influenza vaccines contributes to increased vaccination uptake.^22^ This approach helps reduce the burden of individually mailing COVID-19 vaccination screening questionnaires.

Regarding sociodemographic factors, COVID-19 vaccination uptake was negatively associated with ADI and positively associated with municipality-level social economic status. Socioeconomic status is a critical determinant of health behaviors, including vaccination.^23–25^ Previous studies on regional disparities in vaccination coverage have also reported area-level socioeconomic status as factor contributing to regional disparities.^9,24,26^ This indicates that vaccination uptake tends to be lower in municipalities with high deprivation levels, and it is important to implement vaccination policies that reduce the regional disparities. By contrast, the positive association with a higher proportion of the population aged 75 and older may reflect several factors, as follows. Individuals aged 75 years and over might perceive greater personal risk from COVID-19, or they may be more regularly engaged with the healthcare system. A previous study reported that vaccination behavior could also be driven by the desire to avoid becoming a burden on families.^27^ In addition, recommendations from family physicians have been reported to lead to behavioral changes in vaccine uptake among older adults.^28^

These factors may make individuals aged 75 years and older more likely to receive COVID-19 vaccination. In this study, the proportion of workers aged 65 and older was negatively associated with vaccination uptake. This may be due to healthy worker bias. This means that people who work tend to be healthier than those who do not and may be less concerned about the risks of COVID-19 and, thus, less likely to receive vaccinations.

To improve vaccination uptake, it is necessary to consider a multicomponent approach.^29,30^ This study identified potentially effective interventions for improving COVID-19 vaccination uptake. Ensuring low out-of-pocket expenses is a fundamental requirement, and our study suggests that proactive individual notifications are a particularly effective strategy for encouraging vaccination. This approach is not only a means of conveying information, but is also a tool for ensuring equity in access to vaccination opportunities, especially for older adults or for those who are passive about their health. Unfortunately, national funds for COVID-19 vaccinations are scheduled to be abolished by the 2025–2026 season.^31^ To effectively increase vaccination uptake, municipalities may consider continuing financial support, mailing individual notifications, and targeting interventions to specific groups with potentially low vaccination uptake, such as those with low socioeconomic status.

This study has several limitations. First, based on COVID-19 vaccine production, inventory, and return in Japan, the outcome variable—the number of vaccinated individuals—was estimated from vaccine distribution data and not from actual administration records. Furthermore, this study may have been affected by bias if the choice of vaccine manufacturer varied systematically across municipalities.

Second, some variables relied on the most recent but not concurrent data, such as sociodemographic information from the 2020 census, and policy data collected from websites may not have captured all the implemented initiatives. Third, individual-level factors such as personal health beliefs and vaccination intentions were not considered. Finally, this was an ecological study susceptible to ecological fallacy. Despite these limitations, this study provides the first nationwide evidence of the municipal-level drivers of COVID-19 vaccination uptake in the new routine immunization phase, offering valuable insights for policymakers.

## Conclusions

The COVID-19 vaccination uptake varied greatly among municipalities. This study identified several sociodemographic factors and municipal policies associated with COVID-19 vaccination uptake among older adults in Japan’s routine immunization programs. Mailing vaccine screening questionnaires and providing financial support may be effective strategies to improve the COVID-19 vaccination uptake. Policies targeting populations with potentially low vaccination uptake, such as those with a low socioeconomic status, may also be necessary. With an anticipated reduction in government support, the effective implementation of vaccination programs by municipalities will be important to increase COVID-19 vaccination uptake.

## Author contribution statement

Masaki Machida: Conceptualization, Methodology, Formal analysis, Data curation, Writing – original draft, Writing – review & editing.

Akira Nomachi: Conceptualization, Methodology, Formal analysis, Data curation, Writing – original draft, Writing – review & editing.

Kosuke Tanabe: Conceptualization, Investigation, Data curation, Writing – review & editing, Project administration.

Shuhei Ito: Conceptualization, Investigation, Data curation, Writing – review & editing, Project administration, Supervision.

Tomoki Nakaya: Methodology, Resources, Writing – review & editing.

Takashi Matono: Conceptualization, Methodology, Data curation, Writing – original draft, Writing – review & editing.

All authors have read and approved the final manuscript.

## Data Availability

All data produced in the present study are unavailable.

## Acknowledgments

We would like to thank the Fuji Keizai Group for the survey of municipal policies, the Clinical Study Support for the statistical analysis, and Editage for English language editing.

## Data availability

The authors confirm that all data used in the analysis are disclosed within the article. Sociodemographic data are from public domain sources (https://www.e-stat.go.jp/en).

## Author biographies

Masaki Machida is an Associate Professor at Tokyo Medical University, specializing in epidemiology and vaccine hesitancy.

Akira Nomachi specializes in vaccines for infectious disease.

